# Development of a search filter to retrieve reports of interrupted time series studies from MEDLINE and PubMed

**DOI:** 10.1101/2023.09.14.23295594

**Authors:** Phi-Yen Nguyen, Joanne E. McKenzie, Simon L. Turner, Matthew J. Page, Steve McDonald

## Abstract

**Background:** Interrupted time series (ITS) studies contribute importantly to systematic reviews of population-level interventions. However, there is no search filter designed to identify only ITS studies from bibliographic databases. We aimed to develop and validate search filters to retrieve ITS studies in MEDLINE and PubMed.

**Methods:** A set of 1,017 known ITS studies (published 2013-2017) were analysed using text mining to generate candidate terms. A population set of 1,398 time-series studies were used to select discriminating terms. Various combinations of candidate terms were iteratively tested to generate three search filters. An independent set of 700 ITS studies were used to validate the filters’ sensitivities. The filters were test-run in Ovid MEDLINE; their retrieved records were randomly screened for ITS studies to determine their precision. Finally, all three MEDLINE filters were translated to PubMed format and their sensitivities in PubMed were estimated.

**Results:** Three search filters were created: a *precision-maximising* filter with high precision (78%; 95% CI 74%-82%) but moderate sensitivity (63%; 59%-66%), useful for rapid reviews; a *sensitivity-and-precision-maximising* filter with high sensitivity (81%; 77%-83%) but lower precision (32%; 28%-36%), useful when a balance of expediency and comprehensiveness is required; and a s*ensitivity-maximising* filter with high sensitivity (88%; 85%-90%) but likely very low precision, useful for search strategies accompanying specific content terms.

**Conclusion:** Our filters strike different balances between comprehensiveness and screening workload and suit different research needs. To improve retrieval of ITS studies, authors need to identify ITS designs in titles more frequently.

## INTRODUCTION

An interrupted time series (ITS) is a non-randomised design commonly used to evaluate the effects of health policies or natural events. In an ITS study, multiple measurements are collected at (generally) regular time intervals before and after an ‘interruption’. The effect of the interruption can be estimated by comparing pre- and post-interruption data, while controlling for the underlying trend of the pre-interruption period and other time-varying confounding effects such as seasonality ^1^. The ITS design is thus ideal to examine the effects of policy interventions targeted at populations, when randomised trials are impractical or unethical (e.g. a policy restricting alcohol sale to reduce alcohol-related mortality) ^2–4^. Moreover, ITS studies also enable analysis of existing longitudinal administrative data and retrospective evaluation of interventions that were implemented without randomisation ^5,6^.

In systematic reviews examining the effects of policy and health system interventions, ITS studies are likely to be an eligible design, especially when evidence from randomised trials is limited or not available ^7^. The validity of the findings from such reviews rests on the ability to locate all (or most) of the eligible ITS studies. Furthermore, locating ITS studies is also important in the context of meta-research studies, which aim to examine design features, analysis methods, and the quality of reporting of ITS studies ^2–4,8–11^. Such investigations are needed to uncover gaps in methods and underpin the need for methodological research and guidance. For these reasons, developing search filters to identify ITS studies from bibliographic databases is valuable for systematic reviewers and meta-researchers.

There are single search filters designed to locate multiple non-randomised designs (e.g. cohort studies, controlled before-after studies, ITS studies) ^12,13^. However, these do not include terms specific to the ITS design and their performance has not been evaluated against a set of ITS studies. Ad hoc search filters have also been developed in the context of methodological studies ^1– 4,6,8–11^ and systematic reviews of ITS studies ^14,15^; however, the performance of these has not been evaluated. Therefore, in this study, we aimed to develop and examine the performance of search filters to retrieve ITS studies in MEDLINE and PubMed. These databases were chosen because they are more likely to include ITS studies examining the effects of policy and health system interventions compared with other databases.

## METHODS

### Overview

Our approach (Figure 1) was based on the previous studies of Lunny et al. ^16^ and Taljaard et al. ^17^. In brief, a set of known ITS studies published between 2013-2017 was analysed using text mining to generate a list of potential candidate search terms (Steps 1 and 2). We tested these terms against a population set of (non-interrupted) time series studies to identify terms that can discriminate between ITS and time series studies (Step 2). After iteratively testing various combinations of selected terms, we formulated three search filters for the Ovid MEDLINE interface: a *precision-maximising* filter, a *sensitivity-and-precision-maximising* filter and a *sensitivity-maximising* filter (Step 3). All filters were tested against an independent set of ITS studies to validate their sensitivity (Step 4). We ran the *precision-maximising* and *sensitivity-and-precision-maximising* filters in Ovid MEDLINE to retrieve ITS studies indexed in 2018-2021, and estimated their precision. Finally, all three Ovid MEDLINE filters were translated to the PubMed format and evaluated for their sensitivity on the PubMed interface (Step 5). We prepared a protocol for the study (unpublished); all deviations from the protocol are outlined in Supplementary File S1.

**Figure 1.**
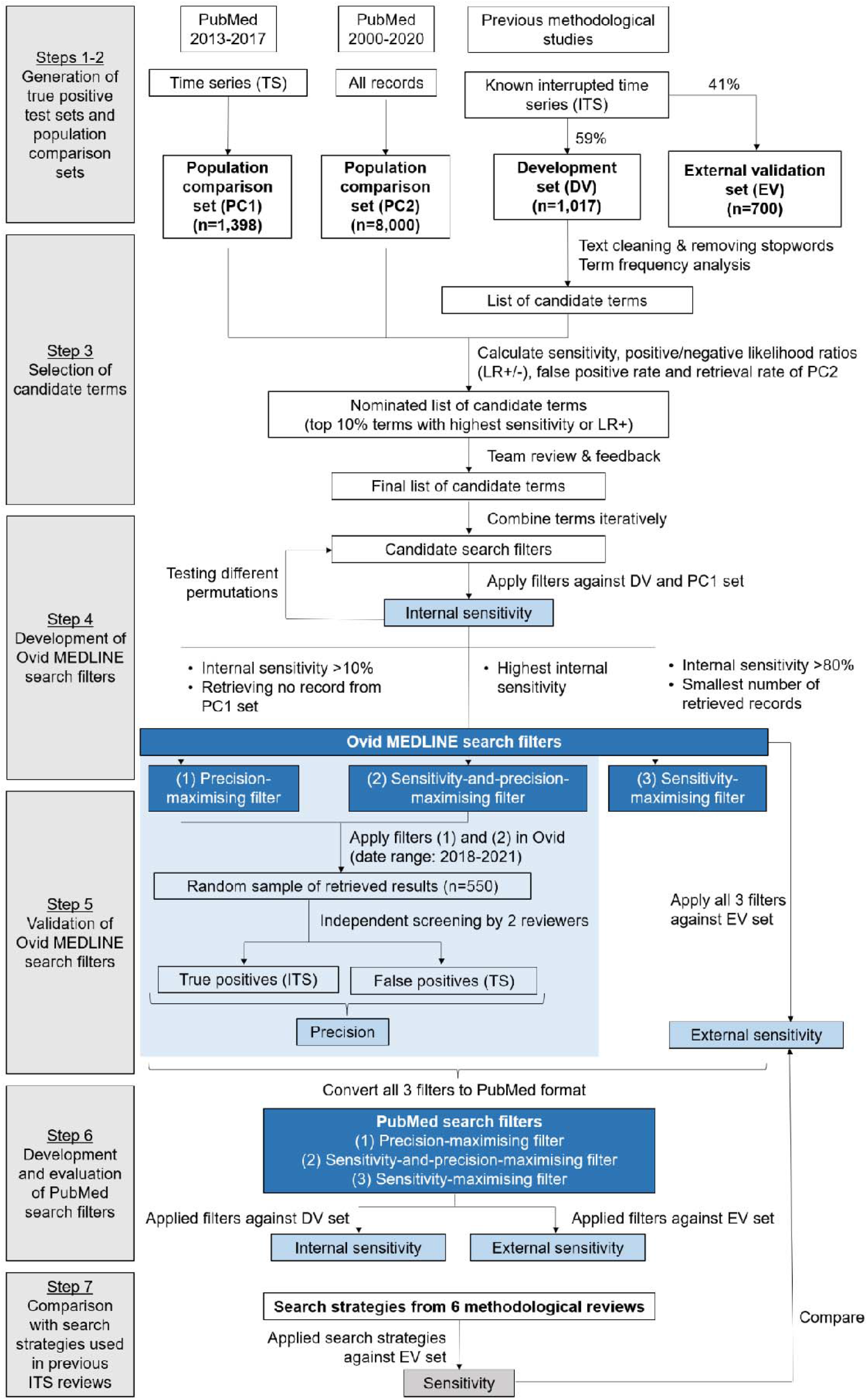
Flow chart summarising the process of developing the search filters.

### Step 1. Generation of true positive test sets

We extracted the lists of included studies from nine methodological reviews of ITS studies ^1–4,6,8–11^. The PubMed records of these studies were imported to EndNote, where duplicates were removed. Among the remaining unique records, we randomly allocated 700 records to an external validation (EV) set and used the remaining records to form a development (DV) set. The DV set was analysed using text frequency analysis to generate a list of words or phrases frequently used in reports of ITS studies. The EV set was an independent set of records used to externally evaluate the sensitivity of the final search filters. The sample size of 700 for the EV set was chosen to yield a 95% confidence interval width of ±3.0%, assuming an underlying true sensitivity of 80%.

### Step 2. Generation of population comparison (control) sets

Two population control sets were created. The first set (PC1) consisted of time series studies indexed in PubMed between 2013-2017, retrieved using a search strategy that searches for time series studies but excludes studies highly likely to be ITS studies (Supplementary File S2). Two authors (PN and ST) independently screened abstracts/titles and full texts of these studies based on pre-defined criteria (Supplementary File S2). Only studies confirmed as time series studies were included in this population set. The second set (PC2) consisted of a random sample of 8,000 PubMed records indexed between 2000-2020 (Supplementary File S2). This sample was designed to represent the general health literature as found in PubMed, allowing us to compare the number of records captured by different search terms. A sample of this size was chosen to allow retrieval rate to be estimated to within (at worst) ±1%, assuming an underlying true rate of 80%.

### Step 3. Selection of candidate terms

Using the *quanteda* package v3.2.1 in R v4.2.0, we performed text frequency analysis on the abstracts and titles from the DV set to identify the words and phrases most frequently used in the reporting of ITS studies (hereafter called the ‘candidate terms’). Prior to performing this analysis, we removed numbers, punctuations and stop-words (e.g. *the, and, he/she*) and stemmed each word to obtain the root words, using the *snowball* lexicon ^18^. Frequency counts were obtained for single words (unigrams), two consecutive words (bigrams) and three consecutive words (trigrams).

A second text frequency analysis was performed on a subset of studies that did not contain the word *interrupted* in their titles. If a word or phrase had a relatively higher frequency in this subset, it was also considered for the candidate term list. This ensured that we captured relevant terms among studies not labelled *interrupted time series* in their titles.

For each candidate term, we tested how well it differentiated between records in the DV set and the PC sets using a range of parameters (Table 1) including: (a) sensitivity, defined as the percentage of records in the DV set that included the term; (b) false positive rate (FPR) of the PC1 set (i.e. population control set containing time series studies), defined as the percentage of records in the PC1 set that included the term; and (c) positive likelihood ratio (LR+), defined as sensitivity divided by FPR. Sensitivity measures the ability of the candidate term to accurately identify ITS studies. False positive rate measures the probability that the candidate term inaccurately identifies time series studies as ITS studies. The positive likelihood ratio measures the ability of a candidate term to differentiate between ITS and time series, with values much greater than 1 indicating that the probability of a true positive exceeds the probability of a false positive. Finally, we calculated the retrieval rate of the PC2 set (i.e. population control set containing random records from PubMed), defined as the number of records in the PC2 set that included the term. The PC2 retrieval rate measures the precision of the candidate term when searching in PubMed.

**Table 1.** Formulas for calculating the performance parameters.

| Parameter | Formula | Interpretation |
| --- | --- | --- |
| **For each candidate term** |  |  |
| Sensitivity | $\frac{\text{No. of records in DV set retrieved by term}}{\text{Total no. of records in DV set}}$ | Higher percentages indicate a higher probability that the term correctly identifies an ITS study. |
| False positive rate | $\frac{\text{No. of records in PC1 set retrieved by term}}{\text{Total no. of records in PC1 set}}$ | Higher percentages indicate a higher probability that the term incorrectly identifies a time series study as an ITS study. |
| Positive likelihood ratio | $\frac{\text{Sensitivity}}{\text{False positive rate}}$ | Measures the ability of a candidate term to differentiate between ITS and time series, with values >1 indicating that the probability of a true positive exceeds the probability of a false positive. |
| Retrieval rate of PC2 set | $\frac{\text{No. of records in PC2 set retrieved by term}}{\text{Total no. of records in PC2 set}}$ | Higher percentages indicate a larger number of records retrieved from general literature. |
| **For search filter** |  |  |
| Internal sensitivity | $\frac{\text{Number of DV records retrieved by filter}}{\text{Total number of records in DV set}}$ | Higher percentages indicate a higher probability that the term correctly retrieves an ITS study. |
| External sensitivity | $\frac{\text{Number of EV records retrieved by filter}}{\text{Total number of records in EV set}}$ | |
| Precision | $\frac{\text{Number of ITS studies retrieved by filter}}{\text{Total number of records retrieved by filter}}$ | Higher percentages indicate a higher probability that a record retrieved by the filter is a true ITS study. |

*Abbreviation: DV: development set; EV: evaluation set; ITS: interrupted time series; PC1: population control set 1 (containing time series studies); PC2: population control set 2 (containing random records from PubMed)*

**Table 2.** Search filters.

| Ovid MEDLINE interface |  |
| --- | --- |
| Precision-maximising | **Terms highly specific to ITS studies**<br>Interrupted Time Series Analysis/ OR (interrupt* time* series OR (segment\$2 adj3 regression)).tw,kf. |
| Sensitivity-and-precision-maximising | **Terms highly specific to ITS studies**<br>1. Interrupted Time Series Analysis/ OR (interrupt* time* series OR (segment\$2 adj3 regression)).tw,kf.<br>**Terms specific to ITS studies**<br>2. (integrat* moving average or slope change or ((piecewise or piece-wise) adj3 regression)).tw,kf.<br>**Combining *time\* series* with other terms not specific to ITS studies**<br>3. time* series.tw,kf.<br>4. (((pre or before or prior) adj5 (post or after or follow*)) or quasi-experiment* or quasiexperiment* or natural experiment* or ARIMA or autoregress* or auto-regress* or segmented or segments or piecewise or piece-wise or interrupt* or implement* or guideline* or prescription* or prescribe* or stewardship or rates or intervention).tw,kf.<br>5. 1 OR 2 OR (3 AND 4) |
| Sensitivity-maximising | **Terms highly specific to ITS studies**<br>1. Interrupted Time Series Analysis/ OR (segment\$2 adj3 regression).tw,kf.<br>**Terms specific to ITS studies and the term *time\* series***<br>2. (time* series or slope change or ((piecewise or piece-wise) adj3 regression)).tw,kf.<br>**Terms not specific to ITS studies**<br>3. (implement* OR rates).tw,kf.<br>4. (((pre OR before OR prior) adj5 (post OR after OR follow*)) OR quasi-experiment* OR quasiexperiment* OR natural experiment* OR ARIMA OR autoregress* OR auto-regress* OR integrat* moving average OR segmented OR segments OR piecewise OR piece-wise).tw,kf.<br>5. 1 OR 2 OR (3 AND 4) |

| PubMed interface |  |
| --- | --- |
| Precision-maximising | Interrupted Time Series Analysis[Mesh] OR interrupted time series[tiab] OR interrupted times series[tiab] OR "segmented regression"[tiab:~3] |
| Sensitivity-and-precision-maximising | **Line-by-line search**<br>#1 Interrupted Time Series Analysis[Mesh] OR interrupted time series[tiab] OR interrupted times series[tiab] OR "segmented regression"[tiab:~3]<br>#2 integrat* moving average[tiab] OR slope change[tiab] OR "piecewise regression"[tiab:~3] OR "piece-wise regression"[tiab:~3]<br>#3 time series[tiab] OR times series[tiab]<br>#4 "pre post"[tiab:~5] OR "before after"[tiab:~5] OR quasi-experiment*[tiab] OR quasiexperiment*[tiab] OR natural experiment*[tiab] OR ARIMA[tiab] OR autoregress*[tiab] OR auto-regress*[tiab] OR segmented[tiab] OR segments[tiab] OR piecewise[tiab] OR piece-wise[tiab] OR interrupt*[tiab] OR implement*[tiab] OR guideline*[tiab] OR prescription*[tiab] OR prescrib*[tiab] OR stewardship[tiab] OR rates[tiab] OR intervention[tiab]<br>#5 #1 OR #2 OR (#3 AND #4) |
|  | **Single line search**<br>Interrupted Time Series Analysis[Mesh] OR interrupted time series[tiab] OR interrupted times series[tiab] OR "segmented regression"[tiab:~3] OR integrat* moving average[tiab] OR slope change[tiab] OR "piecewise regression"[tiab:~3] OR "piece-wise regression"[tiab:~3] OR ((time series[tiab] OR times series[tiab]) AND ("pre post"[tiab:~5] OR "before after"[tiab:~5] OR quasi-experiment*[tiab] OR quasiexperiment*[tiab] OR natural experiment*[tiab] OR ARIMA[tiab] OR autoregress*[tiab] OR auto-regress*[tiab] OR segmented[tiab] OR segments[tiab] OR piecewise[tiab] OR piece-wise[tiab] OR interrupt*[tiab] OR implement*[tiab] OR guideline*[tiab] OR prescription*[tiab] OR prescrib*[tiab] OR stewardship[tiab] OR rates[tiab] OR intervention[tiab])) |
| Sensitivity-maximising | **Advanced search**<br>#1 Interrupted Time Series Analysis[Mesh] OR "segmented regression"[tiab:~3]<br>#2 time series[tiab] OR times series[tiab] OR slope change[tiab] OR "piecewise regression"[tiab:~3] OR "piece-wise regression"[tiab:~3]<br>#3 rates[tiab] OR intervention[tiab]<br>#4 "pre post"[tiab:~5] OR "before after"[tiab:~5] OR quasi-experiment*[tiab] OR quasiexperiment*[tiab] OR natural experiment*[tiab] OR ARIMA[tiab] OR autoregress*[tiab] OR auto-regress*[tiab] OR integrat* moving average[tiab] OR segmented[tiab] OR segments[tiab] OR piecewise[tiab] OR piece-wise[tiab]<br>#5 #1 OR #2 OR (#3 AND #4) |
|  | **Single line search**<br>Interrupted Time Series Analysis[Mesh] OR "segmented regression"[tiab:~3] OR time series[tiab] OR times series[tiab] OR slope change[tiab] OR "piecewise regression"[tiab:~3] OR "piece-wise regression"[tiab:~3] OR ((rates[tiab] OR intervention[tiab]) AND ("pre post"[tiab:~5] OR "before after"[tiab:~5] OR quasi-experiment*[tiab] OR quasiexperiment*[tiab] OR natural experiment*[tiab] OR ARIMA[tiab] OR autoregress*[tiab] OR auto-regress*[tiab] OR integrat* moving average[tiab] OR segmented[tiab] OR segments[tiab] OR piecewise[tiab] OR piece-wise[tiab])) |

A list of potential candidate terms was generated, consisting of the top 10% terms with highest sensitivity or top 10% terms with highest positive likelihood ratios. These threshold values were decided amongst the authors. The list was then reviewed by two authors (JEM and ST) to remove terms not related to study design (e.g. clinical terms). All authors then reviewed all parameter estimates together, and in-conjunction considered the usefulness of the terms in practice. We then determined the final list of candidate terms discussion.

#### Step 4. Development of Ovid MEDLINE search filters

Using an iterative process, various permutations of candidate terms were combined to form several candidate search filters. These filters were implemented in Ovid MEDLINE and their internal sensitivities were calculated (Table 1). After testing, we selected three final search filters that would serve different purposes for users by striking different balances in terms of comprehensiveness and screening workload. The *precision-maximising* filter was designed to achieve the highest internal sensitivity while maintaining an FPR of 0%, so that it has the best differentiating power between ITS and time series studies. The *sensitivity-and-precision-maximising* filter was designed to retrieve the smallest number of records while maintaining an internal sensitivity of at least 80%, to strike a balance between sensitivity (i.e. comprehensiveness) and precision (i.e. less screening workload). The *sensitivity-maximising* filter was designed to achieve the highest internal sensitivity, without consideration for the number of records retrieved nor the FPR (i.e. comprehensive, but with a potentially high screening workload.

#### Step 5. Validation of Ovid MEDLINE search filters for sensitivity and precision

All three filters were tested against an external validation (EV) set of ITS studies (independent of the DV set). For each filter, we calculated their external sensitivities (Table 1). In addition, we assessed the precision of the *precision-maximising* and the *sensitivity-and-precision-maximising* filters (Table 1). To achieve this, we implemented these search filters in Ovid MEDLINE for the period of 2018-2021. From each filter’s results, we randomly sampled 550 records, ensuring that the samples were non-overlapping. These records were then screened independently by two authors (PN and ST) and classified as an ITS (and thus a true positive) if the author described the study as an ITS study, or if the study had the following characteristics: (a) time series data with at least two segments separated by a clearly defined intervention or exposure, (b) observations collected on a group of individuals (e.g., community, hospital) at each time point, and (c) three or more data points per segment for at least two segments (full details in Supplementary File S3). Any discrepancy during screening was resolved by discussion between PN and ST, or by referral to JEM. A sample of 550 allowed the precision to be estimated to within ±2.5% (assuming a true underlying precision of 10%). The precision was calculated for filters (Table 1). Note that we did not estimate the precision of the *sensitivity-maximising* filter, as maximising precision was not the focus of this filter.

#### Step 6. Development and evaluation of PubMed search filters

All three filters were converted to PubMed format, preserving the search fields (title/abstract versus keywords) and proximity search operators, where possible, so that both PubMed and Ovid MEDLINE versions were comparable. All three PubMed filters were applied against the DV and EV sets to evaluate their internal and external sensitivity (Table 1).

#### Step 7. Comparison with search strategies used in previous ITS reviews

We examined the performance of six (of nine) of the search strategies presented in the methodological reviews that formed the DV set ^2–4,8,9,19^ (Supplemental File S4). The other methodological reviews did not provide a search strategy ^1^ or did not use a search strategy ^6,10^. Performance was examined by running these search strategies on the PC1 and EV sets and calculating their sensitivities and positive likelihood ratios (Table 1). These performance parameters were compared with those of our search filters.

## RESULTS

### Step 1. Generation of true positive test sets

From nine methodological reviews of ITS studies ^1–4,6,8–11^, a total of 2,144 ITS studies were identified. After removing duplicates and studies without PubMed abstracts, the final sample consisted of 1,717 ITS studies. After randomly allocating 700 records (41%) to the EV set, the remaining DV set consisted of 1,017 records (59%).

### Step 2. Generation of population comparison (control) sets

The search strategy to locate time-series studies (Supplementary File S2, section 1) yielded a total of 4,198 records indexed between 2013-2017 from PubMed. Of these, 2,440 records did not meet our inclusion criteria (Supplementary File S2, section 2) at title/abstract screening, and a further 360 studies were excluded at full-text screening. This resulted in 1,398 studies identified as time-series studies, which formed the PC1 set.

### Step 3. Selection of candidate terms

After team review and consensus, 42 candidate terms were shortlisted, consisting of one MeSH heading and 41 phrases to be searched as text words (.tw.) or keyword heading words (.kf.) ^20^ (Supplementary File S5). The candidate terms were categorised into three broad groups: terms found to be highly specific (n=3), specific (n=11), and not specific to the ITS study design (n=28), with these categorisations based on estimates of sensitivity, FPR and the number of records retrieved. *Highly specific* terms were those that had an FPR of 0% and a sensitivity of more than 10% (range 15% to 53%). This group included the phrases *interrupted time series* and *segmented regression*, a regression technique commonly used in ITS studies to estimate and compare pre- and post-intervention trends ^1,21^. *Specific* terms were those that had a small FPR (≤1%) and retrieved a small number of records (≤15,000) across the whole of Ovid MEDLINE from 2000-2020. Their sensitivity estimates were less than 10% (range <0.5% to 9%). This group included other descriptors of statistical methods (e.g. autoregressive integrated moving average or piece-wise regression) and non-randomised study designs (e.g. quasi-experiments or natural experiments). *Non-specific* terms were those that had a low sensitivity (≤10%), large FPR (>1%) or retrieved a large number of records (>15,000) in Ovid MEDLINE. This group included terms describing types of interventions (e.g. quality improvement, policy or programme interventions), effect metrics (e.g. slope change), and terms suggesting time segmentation (e.g. pre/post, before/after, intervals or segments).

### Step 4. Development of Ovid MEDLINE search filters

Terms found to be highly specific to the ITS study design were combined to form the first filter (*precision-maximising* filter). We formed a second filter (*sensitivity-and-precision-maximising* filter) by adding two components to the first filter: (a) terms found to be specific to the ITS study design, and (b) a combination of the phrase *time* series* and terms found to be not specific to ITS studies. The term *time* series* had high sensitivity and low precision: searching using this term yielded only 74% (n=755) of the ITS studies in the DV set, but 100% of the time series studies in the PC1 set (Supplementary File S5). However, combining this phrase with non-specific terms struck a balance between sensitivity and precision, meeting the intended purpose of this filter. The third filter (*sensitivity-maximising* filter) was constructed by combining terms from all three groups (highly specific, specific and non-specific terms). When evaluated against the DV set, our filters internal sensitivities were estimated as 64% (95% CI 61% to 67%) (*precision-maximising*), 83% (95% CI 81% to 86%) (*sensitivity-and-precision-maximising*) and 88% (95% CI 86% to 90%) (*sensitivity-maximising*). The *sensitivity-and-precision-maximising* search filter retrieved over a third (36%) of the time series studies, and the *sensitivity-maximising* search filter retrieved all of the time series studies (Table 3).

**Table 3.** Evaluation of search filters.

| Name of filter | Total records retrieved between 2000-2020 | No. of ITS from DV set retrieved (N=1,017) |  | No. of time series from PC1 retrieved (N=1,398) |  | No. of ITS from EV set retrieved (N=700) |  | No. of ITS from precision test set retrieved (N=550) |  |
| --- | --- | --- | --- | --- | --- | --- | --- | --- | --- |
|  |  | n | Internal sensitivity | n | False positive rate | n | External sensitivity | n | Precision |
| Ovid MEDLINE interface |  |  |  |  |  |  |  |  |  |
| Precision-maximising filter | 4,392 | 654 | 64%<br>(61% to 67%) | 0 | 0%<br>(0% to 0.3%) | 438 | 63%<br>(59% to 66%) | 430 | 78%<br>(74% to 82%) |
| Sensitivity-and-precision-maximising filter | 13,078 | 848 | 83%<br>(81% to 86%) | 503 | 36%<br>(33% to 39%) | 564 | 81%<br>(77% to 83%) | 177 | 32%<br>(28% to 36%) |
| Sensitivity-maximising filter | 90,944 | 896 | 88%<br>(86% to 90%) | 1398 | 100%<br>(99.7% to 100%) | 616 | 88%<br>(85% to 90%) | - |  |
| PubMed interface |  |  |  |  |  |  |  |  |  |
| Precision-maximising filter | 4,299 | 652 | 64%<br>(61% to 67%) | - |  | 435 | 62%<br>(58% to 66%) | - |  |
| Sensitivity-and-precision-maximising filter | 13,254 | 848 | 83%<br>(81% to 86%) |  |  | 562 | 80%<br>(77% to 83%) |  |  |
| Sensitivity-maximising filter | 114,869 | 893 | 88%<br>(86% to 90%) |  |  | 608 | 87%<br>(84% to 89%) |  |  |

Abbreviation: DV: development set; EV: evaluation set; PC1: population control set 1 (containing time series studies); ITS: interrupted time series studies
Notes: 95% confidence intervals for the percentages were calculated using the exact binomial method.

### Step 5. Validation of the Ovid MEDLINE search filters for sensitivity and precision

Our search filters demonstrated external sensitivities of 63% (95% CI 59% to 66%) (*precision-maximising*), 81% (95% CI 77% to 83%) (*sensitivity-and-precision-maximising*) and 88% (95% CI 85% to 90%) (*sensitivity-maximising*). These estimated external sensitivities were similar to the internal sensitivities. The precision was estimated as 78% (95% CI 74% to 82%) and 32% (95% CI 28% to 36%) for the *precision-maximising* and *sensitivity-and-precision-maximising* filters, respectively (Table 3). Compared with the *sensitivity-and-precision-maximising* filter, the *sensitivity-maximising* filter increased the external sensitivity by 5 percentage points, but retrieved 77,800 more records (Table 4).

**Table 4.** Comparison of performance with search strategies used in other methodological studies.

| Name/source of filter |  | Total records retrieved (2000-2020) | Sensitivity based on evaluation set | False positive rate |
| --- | --- | --- | --- | --- |
| *Precision-maximising filter* | Ovid MEDLINE | 4,392 | 63% (59% to 66%) | 0% (0% to 0.3%) |
|  | PubMed | 4,299 | 62% (58% to 66%) | - |
| *Sensitivity-and-precision-maximising filter* | Ovid MEDLINE | 13,078 | 81% (77% to 83%) | 36% (33% to 39%) |
|  | PubMed | 13,254 | 80% (77% to 83%) | - |
| *Sensitivity-maximising filter* | Ovid MEDLINE | 90,944 | 88% (85% to 90%) | 100% (99.7% to 100%) |
|  | PubMed | 114,869 | 87% (84% to 89%) | - |
| Ewusie 2020 |  | 5,089 | 67% (64% to 71%) | 6% (5% to 7%) |
| Hategeka 2020 |  | 25,889 | 77% (74% to 80%) | 100% (99.7% to 100%) |
| Hudson 2019 |  | 41,393 | 66% (62% to 70%) | 3% (2% to 4%) |
| Jandoc 2015 |  | 4,666 | 32% (29% to 36%) | 8% (7% to 10%) |
| Korevaar 2022 |  | 78,029 | 82% (79% to 85%) | 100% (99.7% to 100%) |
| Turner 2020 |  | 8,280 | 65% (62% to 69%) | 0.1% (0% to 0.5%) |

*Notes: 95% confidence intervals for the percentages were calculated using the exact binomial method.*

### Step 6. Evaluation of PubMed search filters

The Ovid MEDLINE filters were translated into PubMed syntax with minimal changes, except for some terms which were simplified since the proximity search function in PubMed does not allow complex Boolean combinations or truncation ^22^. Overall, the PubMed versions showed similar performance to the Ovid MEDLINE versions. In PubMed, the three filters demonstrated internal sensitivities of 64% (95% CI 61% to 67%) (*precision-maximising* filter), 83% (95% CI 81% to 86%) (*sensitivity-and-precision-maximising* filter) and 88% (95% CI 86% to 90%) (*sensitivity-maximising* filter). Their external sensitivities were 62% (95% CI 58% to 66%) (*precision-maximising* filter), 80% (95% CI 77% to 83%) (*sensitivity-and-precision-maximising* filter) and 87% (95% CI 84% to 89%) (*sensitivity-maximising* filter). The total number of records retrieved was similar to those of the Ovid MEDLINE filter, except for the *sensitivity-maximising* filter, which retrieved 26% more records in PubMed than in Ovid MEDLINE for the period of 2000-2020 (Table 3).

### Step 7. Comparison with search strategies used in previous ITS reviews

The search strategies from six other methodological reviews ^2–4,8,9,19^ had estimated sensitivities ranging from 32% (95% CI 29% to 36%) to 82% (95% CI 79% to 85%). Two search strategies ^3,9^ had FPR of 100% (95% CI 99.7% to 100%), while the other strategies had FPR ranging from 0.1% (95% CI 0% to 0.5%) to 8% (95% CI 7% to 10%) (Table 4). Compared with these search strategies, our *sensitivity-maximising* filter achieved higher sensitivity (88% in MEDLINE and 87% in PubMed) than all of the comparison search strategies, however it also retrieved more records. Our *sensitivity-and-precision-maximising* filter also achieved higher sensitivity (81% in MEDLINE and 80% in PubMed) than most of the comparison search strategies, and was better at distinguishing between ITS and time series studies (FPR of 36% in MEDLINE) than those with similar sensitivities (FPR of 100%) ^3,9^. Although our Ovid MEDLINE’s *precision-maximising* filter had a lower sensitivity than most of the comparison search strategies (63% in MEDLINE and 62% in PubMed), it had greater ability to distinguish ITS from time series studies (FPR of 0% in MEDLINE), for which there was only one comparable search strategy ^11^.

## DISCUSSION

We developed a set of search filters to retrieve ITS studies for inclusion in systematic reviews and meta-research studies. We developed three filters to serve different purposes: a *precision-maximising* filter, with high precision but moderate sensitivity; a *sensitivity-and-precision-maximising* filter, with high sensitivity but lower precision; and a *sensitivity-maximising* filter, with high sensitivity but likely very low precision. For each filter, we formulated both Ovid MEDLINE and PubMed versions to increase accessibility.

Among the three filters, the *precision-maximising filter* has the best capability to distinguish between ITS and time series studies. It may be most appropriate in the context of rapid reviews, where there is a need to complete a review in a short timeframe with limited resources to screen studies. In such reviews, search strategies that maximise precision over sensitivity may be selected ^23,23^. However, using such a strategy risks missing a large percentage of eligible ITS studies, which should be reported as a limitation of the review. The *sensitivity-and-precision-maximising* filter might be considered for systematic reviews where a balance of expediency and comprehensiveness is required. Finally, the *sensitivity-maximising* filter might be considered for systematic reviews in which the strategy will be appended to specific content terms (e.g. type of intervention), thus reducing the yield of citations, or for meta-research studies in which a random sample of ITS is sought (e.g. Turner et al ^11^).

Our filters are designed for searching bibliographic databases. However, the grey literature is also an important source of publications for evaluations of health systems or health policy research ^24^. Therefore, reviewers collating evidence for such evaluations should supplement bibliographic searches with searches of relevant grey literature sources (e.g. GreyLit, government reports or websites of the organisations commissioning the research) to further identify eligible studies.

Only in approximately 50% of the records in the development set was the ITS design explicitly identified as such. This necessitated the inclusion of other search terms, which were less specific and resulted in increased yields of records. Reporting of the study design in the title or abstract would improve the performance of the search filters and help to ensure that ITS studies are included in systematic reviews. The importance of reporting the study design in the title or abstract of publications has long been recognised, with reporting guidelines for key study designs including an item recommending study authors should do so (e.g. CONSORT for randomised trials ^25^, PRISMA for systematic reviews ^26^). Our results suggest that such an item should be strongly considered for inclusion in a reporting guideline currently under development for ITS studies ^27^.

### Strengths and limitations

To our knowledge, these are the first search filters specifically for ITS studies that were empirically derived from existing ITS studies. The search filters were evaluated using an independent evaluation set, ensuring that the measured sensitivity is applicable outside of the studies used to generate the candidate terms. The sample sizes of the datasets were sufficiently large to accurately estimate the sensitivity and precision.

Our study was not without limitations. First, we did not evaluate the *sensitivity-maximising* filter for precision, since maximising precision was not the purpose of this filter. We instead presented the total number of records retrieved within a fixed period (2000-2020), which can be used as a proxy to compare the levels of precision between the *sensitivity-maximising* filter and the other two filters. Second, the search filters were designed to identify ITS studies based on two criteria only – a time series and an interruption event, which are key characteristics of the ITS design. It is difficult, if not impossible, to develop search filters to identify other characteristics that might be used to define ITS studies (e.g. the number of data points in the pre- and post-interruption segments). Given systematic reviewers may use different criteria for defining an ITS study, using a minimum set of criteria to identify ITS during a search is prudent since this then gives reviewers the flexibility to apply additional criteria during screening. Lastly, even although the selection of candidate terms was guided by the performance parameters of the candidate terms, there was still subjectivity in the decision making and different teams could have made different decisions.

## CONCLUSION

We developed three search filters to identify interrupted time series studies: a *precision-maximising* filter with the ability to distinguish ITS studies from time series studies; a *sensitivity-maximising* filter with high sensitivity but likely very low precision; and a *sensitivity-and-precision-maximising* filter with a balance between sensitivity and precision. We developed versions of these strategies for Ovid MEDLINE and PubMed interfaces. To improve retrieval of ITS studies, and enable their findings to contribute to systematic reviews, study authors need to identify ITS designs in titles and abstracts more frequently.

### HIGHLIGHTS

#### What is already known

- Search filters are an important tool for searching bibliographic databases when conducting systematic reviews, which allows a more focused search and reduces workload during screening.
- Search filters have been developed for various study designs, such as randomised trials or economic evaluations. No search filter has been developed specifically for interrupted time series (ITS) studies.

#### What is new

- We developed three search filters to identify and retrieve ITS studies.
- These filters strike different balances between comprehensiveness and screening workload to suit different research needs.
- The search filters are available for both PubMed and MEDLINE, two major bibliographic databases for public health and medical research.

#### Potential impact for *Research Synthesis Methods* readers

- The search filters will be useful for systematic reviewers and methods researchers who work primarily with ITS studies.
- Researchers can choose from three versions: a *precision-maximising* filter with high precision and moderate sensitivity, suitable for rapid reviews; a *sensitivity-and-precision-maximising* filter with high sensitivity but lower precision, useful when a balance of expediency and comprehensiveness is required; and a s*ensitivity-maximising* filter with high sensitivity but likely low precision, useful for search strategies appended to narrow, specific content terms.

## Data Availability

All data pertaining to the results and conclusion of this paper is available in the main report and the supplementary files. Analytic code and other relevant data files are available at osf.io/gd9x8.

