## Supplementary Files for "Development of a search filter to retrieve reports of interrupted time series studies from MEDLINE and PubMed"

### Content page

S1. Deviations from protocol

S2. Creation of population sets PC1 and PC2

S3. Screening criteria for evaluation of precision

S4. Characteristics of data sources

S5. Performance of candidate terms

### S1. Deviations from protocol

| **Original plan** | **Method implemented** | **Reason for modification** |
| --- | --- | --- |
| We planned to calculate the following parameters for each candidate term and each search filter: sensitivity, false positive rate, and positive likelihood ratio. | We also used the total numbers of records retrieved within a fixed time frame (2000-2020) in the selection of candidate terms and final search filters. | Since evaluation of precision was conducted on external sample only after the search filters were finalised, and each evaluation involves screening 550 full texts to confirm the retrieved studies were all ITS, precision was not readily assessable during development of the filter. As the total numbers of records retrieved is inversely related to precision, it was used as a proxy to quickly estimate and compare levels of precision between different permutations of the search filters. |
| We planned to create one search filter, with an additional precision-maximising filter added if the precision of the filter is below 10%. | We created three different search filters with different degrees of sensitivity and precision. | We observed that no single permutation can adequately fulfil all three criteria: maximising sensitivity (to retrieve as many ITS as possible), maximising precision (to reduce time required for screening) and maximising positive likelihood ratios (to reduce numbers of false positives from other time series designs). Therefore, we created three different filters that were tailored to each objective, so that future users can choose the filter best suited for their research purposes. |
| We planned to organise a Delphi-type group discussion to select the list of candidate terms and to review the final search filters, inviting external members who are experts in this research field. | We discussed the list of candidate terms and review the final search filters within the author team. | It was a pragmatic decision due to time and logistic constraints. |
| We did not plan to compare the performance of our search filter against existing search strategies. | We presented and discussed how our search filters compare against search strategies used in six other methodological reviews of ITS studies. | The comparison analysis was initially conducted as part of final review of the search filter, in order to identify areas for improvement. We decided to present the findings for transparency and to add context to our evaluation results. |

### S2. Generation of population sets PC1 and PC2

2.1 **PC1 | PubMed search for 2013-17 records**

**Concept**: time series as phrase in title/abstract less terms that are indicative of ITS studies, limited to Humans, 2013-2017 and Abstracts = 4198 (6 April 2019)

**Syntax**: ("time series"[Title/Abstract]) NOT (("Interrupted Time Series Analysis"[Mesh]) OR (("interrupted time series"[Title/Abstract] OR "change point"[Title/Abstract] OR "segmented regression"[Title/Abstract] OR "segmented linear regression"[Title/Abstract] OR "time-series intervention"[Title/Abstract] OR "phase design"[Title/Abstract] OR "multiple baseline"[Title/Abstract] OR ARIMA[Title/Abstract] OR "integrated moving average"[Title/Abstract]))) Filters: Abstract; Publication date from 2013/01/01 to 2017/12/31; Humans

2.2 **PC1 | Screening criteria**

A study is classified as an ITS study if:

- The author defines the study as ITS design; or
- All of the below:
  - Time series data are provided, featuring at least two segments separated by a clearly defined intervention or exposure; and
  - Observations are collected on a group of individuals (e.g. community, hospital) at each time point; or

A study is classified as a time series study when:

- Time series data are provided, featuring no interruption (i.e. an exposure or an intervention), and
- Observations are collected on individuals or groups of individuals (eg, community, hospital) at each time point.

A study is excluded if it features all other study designs (e.g. single-case designs) or is a methodological paper examining ITS studies.

2.3 **PC2 | Process of obtaining random records from PubMed 2000-2020**

Search query *2000/01/01: 2000/01/01[dp]* was run on PubMed and the PMID of the earliest published record from the results was recorded.

Search query *2020/12/31:2020/12/31[dp]* was run on PubMed and the PMID of the last published record from the results was recorded.

Function RANDBETWEEN() was used in Excel to generate a list of 8,000 random numbers that fall between the two PMIDs above. These PMIDs were applied to PubMed to retrieve 8,000 records for the PC2 set.

### S3. Screening criteria for evaluation of precision

A study is classified as an ITS if:

- The author identified the study as ITS design in the title, abstract or Methods section*^[1]^; or
- All of the below
  - Time series data are provided, featuring at least two segments separated by a clearly defined intervention or exposure; and
  - Three or more data points per segment for at least two segments
  - Observations are collected on a group of individuals (e.g. community, hospital) at each time point; or

The following studies are not classified as ITS:

- Case-based time series i.e. time series that consists of data points from a single individual, without aggregation at a population level
- Studies that do not define the interruption event *a priori* (e.g. studies using join-point analysis to detect points of change in trends)
- Methodological studies examining ITS studies, without using real-world data
- Animal studies

### *S4. Characteristics of data* sources for the development set

**4.1	General characteristics**

| **Source study** | **Description of sample** | **Eligibility criteria / Definition of ITS used in screening** | **Sources searched / sampling process** | **No. of ITS identified / included** | **Date range of the included ITS** | **Search strategy used to retrieve ITS** |
| --- | --- | --- | --- | --- | --- | --- |
| **Bernal, 2018** | Studies of public health interventions, including studies evaluating impact of non-health  interventions on health outcomes | No definition of ITS was provided in the Methods of Methodological Paper 4 (which involves screening for ITS studies). However, authors note the following definition for an ITS in section 3.4.4 - Terminology: “[...] I use the term interrupted time series exclusively for those studies which incorporate trends over time within the model. Studies with a **single pre-intervention** baseline observation and a **single post-intervention** observation or studies in which there are multiple pre- and post-intervention observations but where the time they were measured is **not included** in the model, I regard as simple pre-post or simple before-after designs.” | Database (1): Medline | 105 / 5 | 2014-2015 | No search strategy reported |
| **Ewusie et al, 2020** | Health-related studies that reported on the application of ITS design | Eligibility criteria: Studies with **at least three time points** **before and after** the intervention and had a clearly defined time point or period within which the intervention was implemented were included    No definition of ITS was provided in the Methods. However, authors note the following definition in the Introduction: “With this design, outcomes are measured at different time points before and after implementing an intervention, allowing the change in level and trend of outcomes to be compared, to evaluate intervention effects” | - Databases (7): MEDLINE, JSTOR, PUBMED, EMBASE, CINAHL, Web of Science and the Cochrane Library - Contacting methodological experts for less accessible or unpublished materials | 1,365 / 1,321 | 1978-2018 (96% were published 2000 onwards) | Ovid MEDLINE |
| **Hategeka et al, 2020** | Studies that evaluating health system quality improvement interventions using ITS | Eligibility criteria: ITS with **at least three data points** **before and after** an intervention.    No definition of ITS was provided in the Methods. However, authors note the following definition in the Introduction: “The ITS design relies on data collected at multiple intervals over time (ie, time series data) before and after an intervention to establish a causal relationship between an intervention (eg, QI) and an outcome of interest (eg, health outcomes)”. | - Databases (11): MEDLINE, EMBASE, CINAHL, Web of Science, Global Health, Google Scholar, Africa-Wide, LILACS, IMSEAR, IMEMR, and WRPRIM - Complementing electronic databases - Hand searches of the bibliographies of previous works - Conference proceedings (eg, International Forum on Quality and Safety in Healthcare, Institute of Healthcare Improvement) | 120 / 119 | 1990-2018 (98% were published 2000 onwards) | Ovid MEDLINE |
| **Hudson et al, 2019** | Studies of health or healthcare intervention (e.g. programs, policies, or educational interventions) | Eligibility criteria: ITS with **a minimum of two data points pre and one post-intervention.**    No definition of ITS was provided in the Methods. However, authors note the following definition in the Introduction: “In an ITS design, data are collected at multiple and equally spaced time points (e.g. weekly, monthly, or yearly) before and after an intervention.” | Database (1): Medline | 116 / 98  [18 duplicates] | 2014-2015 | Ovid MEDLINE |
| **Jandoc et al, 2015 ^1^** | Studies of drug utilization in humans | Eligibility criteria: Empirical applications that examined the impact of interventions at the population level.    No definition of ITS was provided in the Methods. Authors note the following definition for an ITS in the Introduction: “Interrupted time series methods use aggregate data collected over equally spaced intervals before and after an intervention, with the key assumption that data trends before the intervention can be extrapolated to predict trends had the intervention not occurred”. | - Databases (1): Medline - Citation search of methodological articles using Web of Science - Hand searches of reference lists from methodological studies | 220 / 216 | 1984 – 2013  (92% were published 2000 onwards) | Ovid MEDLINE  (combined with drug utilization) |
| **Korevaar et al, 2022** | Studies from all disciplines, e.g. public health, psychology, education,  economics | Eligibility criteria: (i) there is a clearly defined timepoint when the interruption occurred; and, 2) **at least three data points** **before and after** the interruption; OR (3) as defined by review author as ITS, regardless of study design | All ITS included in 29 reviews, which in turn were identified by searching the following databases (8): Medline, Embase, Campbell Systematic Reviews, Cochrane Database of Systematic Reviews, 3ie, EconLit, PsycINFO, and ERIC.  Note: As long as a review has at least 2 ITS that met the eligibility criteria, all ITS under that review will be included. | 213 / 132  [7 duplicates] | 1976-2018 (84% were published 2000 onwards) | Ovid MEDLINE |
| **Polus et al, 2017** | Health-related studies from 10 intervention types (behavioural/ educational, clinical, environmental, health policy, health system,  nutrition, occupational, pharmaceutical, screening, and  vaccination interventions) | Eligibility criteria: Studies where authors explicitly used the term ‘‘interrupted time series’’ or “ITS studies” (i.e., merely ‘‘before-after studies’’ or ‘‘time series’’ were excluded).    No definition of ITS was provided in the Methods. Authors note the following definition for an ITS in the Introduction: “An ITS study is defined in the Cochrane Handbook as a study that uses observations at multiple time points before and after an intervention (the ‘‘interruption’’). Cochrane EPOC specifies minimum criteria that ITS studies must use **at least three data points before and three** **after** the intervention and clearly define the point in time when the intervention occurred”. | All ITS included in 16 Cochrane reviews, which in turn were identified by searching the Cochrane Library | 16 / 12 | 2012-2015 | [ITS studies from 16 Cochrane reviews] |
| **Ramsay et al, 2003** | Mass media interventions to improve health  service utilisation, and strategies for clinical guideline  dissemination and implementation | Eligibility criteria: (i) there were **at least three time points before and after** the intervention, irrespective of the statistical analysis used; (ii) the intervention occurred at a clearly defined point in time; and (iii) the study measured provider performance or patient outcome objectively | All ITS studies included the following systematic reviews:  (i) Grimshaw J, Thomas R, MacLennan G, Fraser CR, Ramsay CR, Vale LE, Whitty P, Eccles MP, Matowe L, Shirran L, Wensing MJ. Effectiveness and efficiency of guideline dissemination and implementation strategies.  (ii) Grilli R, Ramsay C, Minozzi S. Mass media interventions: effects on health services utilisation. Cochrane database of systematic reviews. 2002(1). | 58 / 49  [3 duplicates] | 1976 – 1998 | [ITS studies from two SRs] |
| **Turner et al, 2019** | Studies of public health interventions or exposures that have public health implications | Eligibility criteria: (i) there were at least two segments separated by a clearly defined intervention or exposure with at **least three points in each segment**; (2) observations were collected on a group of individuals (e.g., community and hospital) at each time point; OR (3) as defined by author as ITS, regardless of study design | Database (1): PubMed | 200 / 192  [7 duplicates] | 2013 – 2017 | Ovid MEDLINE [converted from PubMed] |

*Note: ITS were included in the development set if they are available in PubMed (with abstracts). Methodological studies of ITS design were excluded.

**4.2	Replicated search filters**

**Ewusie 2020**

| 1 | Interrupted Time Series Analysis.mp. or "change point model$".mp. or Interrupted Time Series Analysis/ or (interrupt$ adj3 (time$ series$ or time-series)).mp. or (interven$ adj3 (time$ series$ or time-series)).mp. or (segment$ adj2 regression$).mp. or ((time$ series$ or time-series) adj2 regression$).mp. |
| --- | --- |

**Hategeka 2020 [combined with “quality improvement”]**

| 1 | Interrupted Time Series Analysis/ or Interrupted Time Series Analysis.mp. or ITS Studies.mp. or Interrupted Time Series.mp. or Trend analys*.mp. or Time trend*.mp. or Time series analys*.mp. or Time series.mp. or Segmented regression.mp. or Piecewise regression.mp. or Broken-stick regression.mp. |
| --- | --- |
| 2 | limit 1 to Humans |

**Hudson 2019**

| 1 | interrupted time series.tw,kw. or (pre adj1 post).tw. or (segmented adj3 regression).tw,kw. or quasi experiment$.tw,kw. or (before adj1 after).tw,kw. or arima.tw,kw. or (trend adj3 analys?s).tw,kw. or (longitudinal adj3 chang$).tw,kw. or autoregressive integrated moving average.tw,kw. |
| --- | --- |
| 2 | limit 1 to Review Articles |
| 3 | randomi$ control$ trial$.ti,tw. or meta analys$.ti,kw. |
| 4 | 1 not (2 or 3) |

**Jandoc 2015 [combined with “drug utilization”]**

| 1 | Time series.tw. or Time trend$.tw. or Trend analys$.tw. or Time series analys$.tw. or Forecast model$.tw. or Intervention analys$.tw. |
| --- | --- |
| 2 | Drug.mp. or Medicat$.mp. or Pharmaceutic$.mp. or Prescri$.mp. or Pharmacoepidemiolog$.mp. or Dispens$.mp. or Drug utili#ation.mp. |
| 3 | 1 and 2 |

**Korevaar 2022**

| 1 | Interrupted Time Series Analysis/ or interrupted time series.mp. or (time series or time trend$ or trend analys?s).mp. or (change point or repeated measures or phase design or multiple baseline$ or difference-in-difference$ or single case research or single case experimental).mp. or (ARIMA or autoregressive integrated moving average or integrated moving average or piecewise regression or segmented regression).mp. |
| --- | --- |

**Turner 2020**

| 1 | Interrupted time series analysis/ or interrupted time series.ti,ab. or change point.ti,ab. or segmented regression.ti,ab. or segmented linear regression.ti,ab. or repeated measures study.ti,ab. or piecewise regression.ti,ab. or time-series intervention.ti,ab. or phase design.ti,ab. or multiple baseline.ti,ab. or ARIMA.ti,ab. or integrated moving average.ti,ab. |
| --- | --- |

### S5. Performance of candidate terms

Candidate terms’ performance was evaluated by searching the terms in the titles, abstracts and keywords of corresponding records in Ovid MEDLINE(R) and Epub Ahead of Print, In-Process, In-Data-Review & Other Non-Indexed Citations, Daily and Versions.

| **Category** | **Term** | **Total records retrieved between  2000-2020** | **No. of ITS from DV set retrieved (N=1,017)** | | **No. of time series from PC1 retrieved (N=1,398)** | | **LR+** |
| --- | --- | --- | --- | --- | --- | --- | --- |
|  |  |  | **n** | **Sensitivity** | **n** | **False positive rate** |  |
| **Terms highly specific to ITS studies (FPR=0% and sensitivity>10%)** | | | | | | | |
|  | Interrupted Time Series Analysis/ | 1,223 | 148 | 15% | 0 | 0% | NA* |
|  | (interrupt* time* series).tw,kf. | 3,311 | 541 | 53% | 0 | 0% | NA |
|  | (segment$2 adj3 regression).tw,kf. | 1,198 | 189 | 19% | 0 | 0% | NA |
| **Terms specific to ITS studies (FPR≤1%, sensitivity****≤10% and numbers of records retrieved≤15,000)** | | | | | | | |
| Effect metrics | (slope change).tw,kf. | 160 | 5 | <0.5% | 0 | 0% | NA |
| Statistical method | (autoregress* or auto-regress*).tw,kf. | 5,839 | 66 | 6% | 58 | 4% | 2 |
|  | ARIMA.tw,kf. | 964 | 50 | 5% | 0 | 0% | NA |
|  | (integrat* moving average).tw,kf. | 974 | 50 | 5% | 1 | <0.5% | 69 |
|  | (piecewise or piece-wise).tw,kf. | 4,114 | 10 | 1% | 8 | 1% | 2 |
|  | ((piecewise or piece-wise) adj3 regression).tw,kf. | 581 | 10 | 1% | 5 | <0.5% | 3 |
| Study design | (natural experiment*).tw,kf. | 2,165 | 17 | 2% | 5 | <0.5% | 5 |
|  | (quasi-experiment* or quasiexperiment*).tw,kf. | 14,034 | 96 | 9% | 4 | <0.5% | 33 |
| **Terms not specific to ITS studies (FPR>1%, sensitivity≤10% or numbers of records retrieved>15,000)** | | | | | | | |
| Presence of time series (multiple time points) | (daily or weekly or monthly or yearly or annually or quarterly).tw,kf. | 544,323 | 387 | 38% | 655 | 47% | 1 |
|  | (every adj2 (day$1 or week$1 or month$1 or year$1)).tw,kf. | 83,476 | 15 | 1% | 21 | 2% | 1 |
|  | (time* series).tw,kf. | 29,916 | 760 | 75% | 1398 | 100% | 1 |
| Time segmentation | (before adj5 after).tw,kf. | 275,163 | 280 | 28% | 16 | 1% | 24 |
|  | (pre adj5 post).tw,kf. | 85,826 | 76 | 7% | 2 | <0.5% | 52 |
|  | ((pre or before or prior) adj5 (post or after or follow*)).tw,kf. | 401,545 | 354 | 35% | 20 | 1% | 24 |
|  | interrupt*.tw,kf. | 48,029 | 548 | 54% | 5 | <0.5% | 151 |
|  | interval$1.tw,kf. | 633,307 | 152 | 15% | 225 | 16% | 1 |
|  | period$1.tw,kf. | 1,028,601 | 476 | 47% | 559 | 40% | 1 |
|  | (segmented or segments).tw,kf. | 108,439 | 203 | 20% | 5 | <0.5% | 56 |
|  | points.tw,kf. | 307,884 | 50 | 5% | 40 | 3% | 2 |
|  | times.tw,kf. | 492,383 | 42 | 4% | 71 | 5% | 1 |
| Intervention type | audit$1.tw,kf. | 30,899 | 41 | 4% | 1 | <0.5% | 56 |
|  | complian*.tw,kf. | 102,308 | 61 | 6% | 4 | <0.5% | 21 |
|  | (decis* support).tw,kf. | 15,307 | 27 | 3% | 3 | <0.5% | 12 |
|  | guideline*.tw,kf. | 322,404 | 124 | 12% | 18 | 1% | 9 |
|  | implement*.tw,kf. | 464,306 | 424 | 42% | 94 | 7% | 6 |
|  | initiative*.tw,kf. | 80,375 | 56 | 6% | 17 | 1% | 5 |
|  | intervention.tw,kf. | 547,314 | 419 | 41% | 40 | 3% | 14 |
|  | (interven* adj2 study).tw,kf. | 22,063 | 33 | 3% | 0 | 0% | NA |
|  | introduc*.tw,kf. | 817,223 | 246 | 24% | 101 | 7% | 3 |
|  | ((patient or clinical) adj outcome*).tw,kf. | 208,632 | 39 | 4% | 4 | <0.5% | 13 |
|  | (policy or policies).tw,kf. | 218,618 | 203 | 20% | 157 | 11% | 2 |
|  | (prescription* or prescrib*).tw,kf. | 184,715 | 279 | 27% | 43 | 3% | 9 |
|  | (prevention or preventive).tw,kf. | 512,665 | 82 | 8% | 126 | 9% | 1 |
|  | project$1.tw,kf. | 167,686 | 29 | 3% | 34 | 2% | 1 |
|  | program$3.tw,kf. | 645,298 | 287 | 28% | 109 | 8% | 4 |
|  | (qualit* adj3 improv*).tw,kf. | 145,714 | 99 | 10% | 19 | 1% | 7 |
|  | stewardship.tw,kf. | 8,295 | 54 | 5% | 4 | <0.5% | 19 |
|  | strateg*.tw,kf. | 1,031,298 | 96 | 9% | 95 | 7% | 1 |
| Other | (interven* adj2 effect*)).tw,kf. | 40,845 | 39 | 4% | 2 | <0.5% | 27 |
|  | rates.tw,kf. | 848,146 | 399 | 39% | 297 | 21% | 2 |

*Abbreviation: DV: development set; EV: evaluation set; LR+: positive likelihood ratio; PC1: population control set 1.*

**NA denotes division by zero, which occurs when no false positive was retrieved.*

1. This is consistent with the underlying principle for search filters (e.g. a search filter for randomised controlled trials (RCT) will search and retrieve any study labelling itself as an RCT), and with the fact that there is no standardised definition for the ITS study design.
